# AI-Based MRI Radiomics for Preoperative Detection of Tumor Deposits in Rectal Cancer: A Bivariate Diagnostic Accuracy Meta-analysis

**DOI:** 10.64898/2026.11.02.26367109

**Authors:** Mohamed A. Alsaied, Mahmoud Hegazy, Maab M. Saleh, Bassant Barakat, Omar Ramadan Ibrahim, Hassan Magdy Abdelrazek, Sedigheh Nazemi, Alaa El-Hussana, Mostafa Shalaby

## Abstract

**Background:** In rectal cancer (RC), tumor deposits (TDs) are clinically important as they are associated with a negative impact on prognosis and affect risk stratification as well as management plans. MRI radiomics combined with artificial intelligence (AI) has been explored for non-invasive preoperative TDs detection. However, results vary across studies; therefore, we performed a systematic review and meta-analysis to pool their diagnostic test accuracy (DTA).

**Methods:** This systematic review and DTA meta-analysis were conducted in accordance with the Cochrane Handbook for Systematic Reviews of Diagnostic Test Accuracy and reported in adherence to the Preferred Reporting Items for Systematic Reviews and Meta-Analyses of Diagnostic Test Accuracy Studies (PRISMA-DTA). Databases were screened for DTA studies of MRI-based radiomics predicting TDs in adult RC patients with histopathology as reference.

**Results:** Nine studies (2589 patients) were identified. Across included studies, the bivariate meta-analysis showed pooled sensitivity of 0.765 (95% CI 0.714 - 0.809) and specificity of 0.790 (95% CI 0.750 - 0.825), with overall AUC of 0.845 and very low heterogeneity (I² = 7.3%). In validation datasets, diagnostic performance remained consistent (AUC 0.806), with pooled sensitivity 0.751 (95% CI 0.681 - 0.809) and specificity 0.751 (95% CI 0.681 - 0.809). For MRI modality, T2 weighted image (T2WI) showed higher sensitivity (0.745) than T2WI plus diffusion weighted image (DWI) (0.697), while T2WI+DWI had higher specificity (0.809 vs 0.737) and AUC (0.829 vs 0.796), with no significant subgroup differences (p≥0.192). Models combining radiomics with clinical/radiological variables achieved significantly higher sensitivity (0.842 vs 0.657, p<0.001) and higher AUC (0.827 vs 0.752), with comparable specificity.

**Conclusion:** MRI-based radiomics demonstrates good accuracy for preoperative detection of tumor deposits in RC. The integration of radiomics with clinical or imaging features increases the accuracy of identification. Amongst the MRI modalities, models that utilize the combination of T2WI plus DWI are more accurate than those using T2WI or DWI only.

## 1 INTRODUCTION

The trends in mortality rates emerged as a better indicator of cancer prognosis rather than incidence or survival which might be affected by screen-detection biases. Colorectal cancer (CRC) emerged as the second leading cause of cancer deaths ^1^ and the first in those younger than 50 years ^2^. Rectal cancer (RC) represents the third most common CRC cases ^1^. Traditionally, the pathological classification of RC is based on tumor depth and lymph node metastasis ^3^. Surprisingly, in 1935, Gabriel et al., ^4^ after pathological dissection of 100 RC specimens, extranodal tumor deposits (ENTDs) were reported without nodal involvement. Later, multiple studies evaluated the significance of ENTDs; however, these studies described ENTDs that do not fit within the TNM or Dukes systems ^5^. Interestingly, the 7^th^ edition of the TNM staging system suggested an N1c subcategory, which was identified as separate malignant cells nodules or deposits in the mesorectal fat, devoid of evidence of residual lymph node tissue ^6^. Recently, the 8^th^ Edition established the classification of ENTDs with the addition of the prerequisites of being devoid of any histological evidence of nodules, vascular, or neural structures ^7^.

The incidence and significance of ENTDs, also known as tumor deposits (TDs), were evaluated by Lord et al., ^8^ in a meta-analysis published in 2017, in which they reported a median prevalence of 21.3% in RC and a significant pooled hazard ratio for adverse overall survival and disease-free survival. Pelvic magnetic resonant imaging (MRI) is now considered one of the pillars for RC staging with a structured reporting checklist including TDs within the mesorectum ^9–11^. However, the role of MRI in identifying TDs remains challenging due to the reliability in differentiating lymph nodes metastasis from TDs and lower sensitivity in detecting TDs less than 5 mm ^12,13^.

Radiomics has emerged as a promising tool which can aid in diagnosis and prognosis of malignancy. It is a risk-free tool capable of extracting and analyzing quantitative features that cannot be captured by the naked eye from conventional medical images through machine learning and artificial intelligence (AI) algorithms, resulting in signatures reflecting tumor phenotype ^14^. Recently, several studies have developed radiomic models incorporating preoperative pelvic MRI to predict TDs in RC patients, resulting in better prediction of TDs. Despite the promising role, the field requires advances to establish optimal feature selection and ideal algorithms. So, in this study, we conduct a systematic review and diagnostic test accuracy (DTA) meta-analysis collating all eligible studies and pooling the paired sensitivity and specificity using a bivariate random-effects model. By isolating workflows that remain accurate across diverse settings, our analysis offers a practical, reproducible radiomics roadmap for routine pre-operative staging of tumor deposits and more informed treatment decisions.

## 2 METHODS

### 2.1 Reporting checklist

We conducted this systematic review and DTA meta-analysis in accordance with the Cochrane Handbook for Systematic Reviews of Diagnostic Test Accuracy ^15^. Reporting of this study adheres to the Preferred Reporting Items for Systematic Reviews and Meta-Analyses of Diagnostic Test Accuracy Studies (PRISMA-DTA) ^16^, the completed checklist is provided in Supplementary Table S1.

### 2.2 Search strategy and information sources

In-depth literature search went through PubMed, the Cochrane Library, SCOPUS, and Web of Science databases. A systematic literature search was employed using these keywords [("rectal neoplasms" OR "rectal cancer" OR "rectal carcinoma" OR "rectal neoplasm*") AND ("magnetic resonance imaging" OR “MRI” OR "MR imaging" OR "magnetic resonance") AND (radiomic* OR "radiomics" OR "texture analysis" OR "feature extraction") up to July 2025 with no limitations or filters to ensure no studies are missed. The choice of broad search terms was a pragmatic decision to include as many studies as possible.

### 2.3 Eligibility criteria

Studies were deemed eligible for inclusion if all the following criteria were met:

***Population:*** Adult patients with histologically confirmed RC who underwent preoperative pelvic MRI.

***Index test:*** MRI-based radiomics features analyzed using AI to detect the presence of TDs.

***Reference test:*** Histopathological assessment of resected specimens as the reference standard for confirming the presence or absence of TDs.

***Study Design:*** Studies of DTA with prospective and retrospective designs including sufficient data to form a 2×2 contingency table; true positives, false positives, false negatives, and true negatives.

***Outcome Measures:*** Reported or permitted the calculation of accuracy metrics including sensitivity and specificity, area under the receiver operating characteristic curve (AUC), or other relevant measures.

Studies were excluded if: 1) the index model did not incorporate MRI-based radiomics or MRI-derived image features (for example, models based solely on clinical variables or on non-MRI imaging modalities such as CT or PET); 2) if the reference standard for TDs was not histopathology of the surgical specimen; 3) if they did not report or did not allow derivation of essential diagnostic accuracy data (including AUC, sensitivity, specificity, or the cell counts required to compute true positives, false positives, false negatives, and true negatives); 4) non-original publications such as reviews, conference abstracts without full data, editorials, letters, expert opinions; 5) special design such as animal studies, phantom studies, and purely technical feasibility studies that did not provide patient-level diagnostic accuracy data; or 6) articles not available in English language.

### 2.4 Study Selection

Initially, the retrieved records identified from previously mentioned databases were imported into EndNote ^17^ to remove duplicates, then the reaming records were exported into the Rayyan platform ^18^ for screening. Two investigators (M.M.S. and M.A.A.) independently screened all records in a two-step process by titles and abstracts followed by full text against the eligibility criteria. Disagreements were resolved by consensus, and when necessary, by consultation with a senior opinion (A.E.).

### 2.5 Data extraction

Data were manually extracted into a Google Spreadsheet by two investigators (M.H. and O.R.I.) and rechecked by a senior investigator (M.S.). Extracted data included baseline characteristics such as the first author’s name and year of publication; study design; setting; country; and imaging modality. Furthermore, the details of segmentation methods, classifier models, feature extraction software, and validation strategies were extracted.

### 2.6 Risk of bias assessment

The quality of the included studies was evaluated using the revised instrument for Quality Assessment of Diagnostic Accuracy Studies 2 (QUADAS-2) tool ^19^ which consists of four domains assessing Risk of bias (patient selection, index test, reference standard and flow of timing). Three investigators (B.B., S.N., and H.M.A.) independently assessed the methodological quality of all included studies. Any disagreements were resolved through discussion, and when necessary, by consultation with senior opinions (A.E. and M.S.). A risk of bias graph was generated using robvis visualization tool ^20^.

### 2.7 Statistical analysis, Subgroups and Heterogeneity

After extracting the data, key diagnostic indicators—sensitivity, specificity, accuracy, and prevalence—were converted into elements of a 2×2 contingency table: true positives (TP), false positives (FP), true negatives (TN), and false negatives (FN) according to the principles of Monaghan et al. 2021 ^21^ and using a preconstructed calculation sheet. From these, confusion matrices were reconstructed for each study included in the review.

A bivariate random-effects model was applied to obtain pooled estimates of sensitivity and specificity and their 95% confidence intervals (CIs), accounting for between-study heterogeneity and the inherent correlation between these two measures ^22^. Forest plots of sensitivity and specificity were generated, showing point estimates from individual studies, pooled estimates, and 95% CIs, thus allowing a more intuitive comparison of variability across the study samples.

To display the overall diagnostic performance and illustrate the trade-off between sensitivity and specificity, a Hierarchical Summary Receiver Operating Characteristic (HSROC) curve was fitted to the parameters produced by the bivariate model ^23^. Within the same framework, the area under the HSROC curve (AUC) was calculated as an aggregate metric of test accuracy. All statistical analyses were conducted on R software (version 4.4.2, 2024) ^24^ using "mada" and "meta" packages.

We stratified studies by MRI input: combined T2-weighted imaging plus diffusion-weighted imaging (T2WI+DWI), T2-weighted imaging alone (T2WI alone), and diffusion-weighted imaging alone (DWI alone); by model structure: radiomics-only models versus radiomics models combined with additional clinico-radiological variables; and by modelling approach: deep learning (DL) versus traditional machine learning (ML) versus logistic regression (LR).

Heterogeneity was measured with the Zhou-and-Dendukuri method, which builds on the classic I² statistic by accounting for the shared correlation between sensitivity and specificity in diagnostic-test-accuracy studies ^25^. The observed percentage was interpreted as low (25%), moderate (50%), or high (75%) variability. If marked differences remained, planned subgroup and sensitivity analyses sought to identify and explain those sources.

## 3 RESULTS

### 3.1 Included studies

A total of 941 records were initially identified through database searching. Ultimately, nine studies with a total 2589 patients (Yang et al. 2021 ^26^, Jin et al. 2023 ^27^, Fu et al. 2023 ^28^, Feng et al. 2023 ^29^, Li et al. 2023 ^30^, Yang et al. 2023 ^31^, Sun et al. 2024 ^32^, Ao et al. 2024 ^33^, and Wang et al. 2025 ^34^) met the preset inclusion criteria and were included in the qualitative review and meta-analysis. **Figure 1** shows the PRISMA flow diagram illustrating the study selection process.

**Figure 1.**
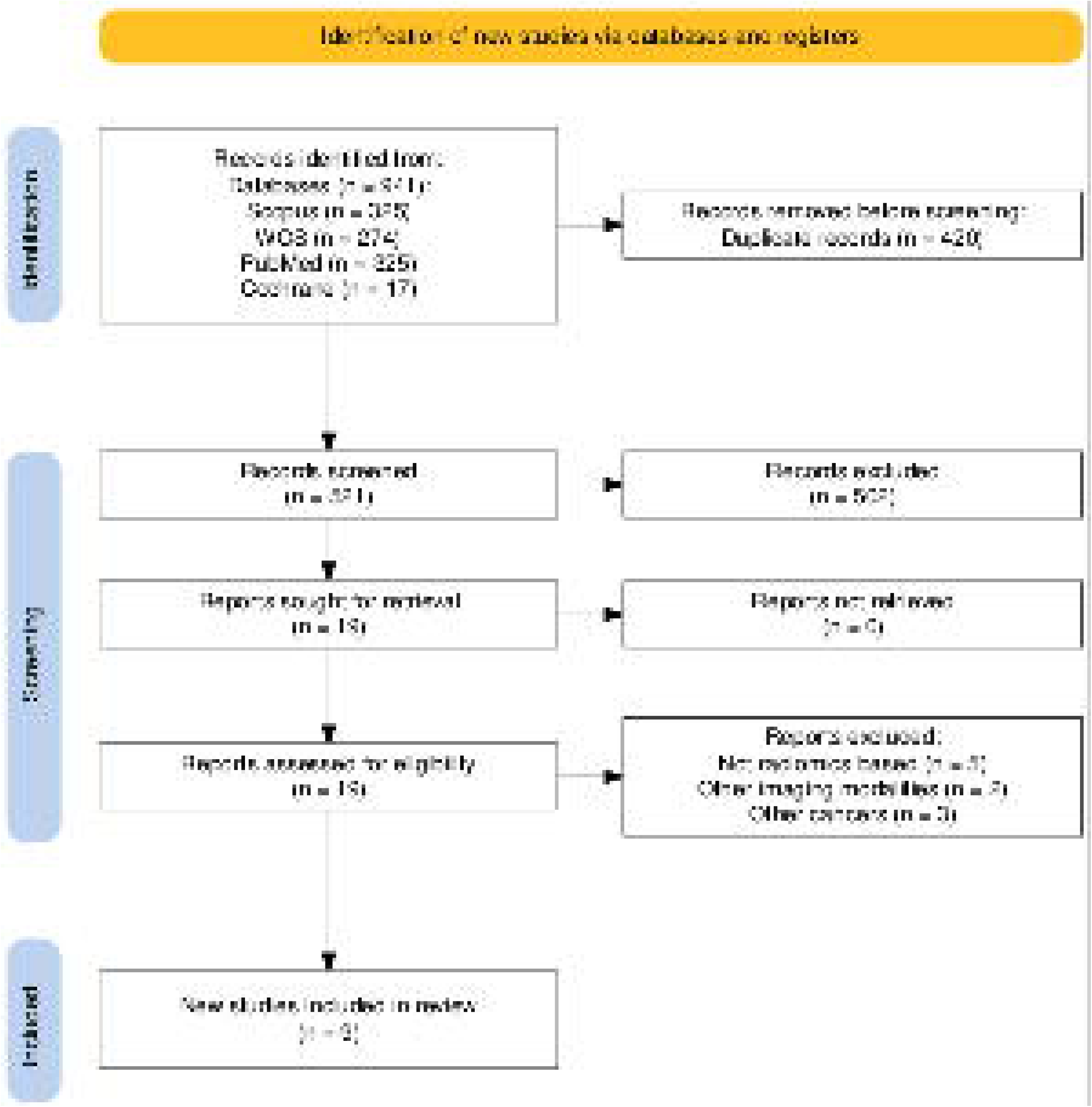
PRISMA 2020 flow diagram of study selection. Flow diagram illustrating the identification, screening, eligibility assessment, and final inclusion of studies retrieved from the searched databases according to PRISMA 2020 guidelines.

All these nine studies were original DTA studies conducted in China between 2021 and 2025. Seven studies were retrospective single-center studies, one was retrospective bicenter designs, and the remaining study was prospective bicenter. They all assessed MRI-based radiomics for the detection of TDs in RC patients, mostly using T2-weighted imaging, frequently in conjunction with diffusion-weighted imaging (DWI), and one with contrast-enhanced T1-weighted imaging (T1C). Classifiers used in the studies included logistic regression (LR) ^26,27,30–34^, least absolute shrinkage and selection operator (LASSO) ^26^, support vector machine (SVM) ^29,32^, random forest (RF) ^32^, multilayer perceptron (MLP) ^28,32^, CatBoost ^34^, XGBoost ^34^, and deep learning models ^28,32^. Feature extraction was done with Pyradiomics ^27–30,34^, 3D Slicer ^31^, AK ^26^, FeAture Explorer Pro (FAE) ^29^, and custom Python and PyTorch scripts ^33^. Manual tumor segmentation was performed in all studies. Validation strategies included k-fold cross-validation (5 or 10-fold) ^26,28,29,32,34^, internal ^33^ or external validation ^27,30,33^, although one study did not report its validation strategy ^31^. **Table 1** shows a detailed summary of the included studies.

**Table 1.**
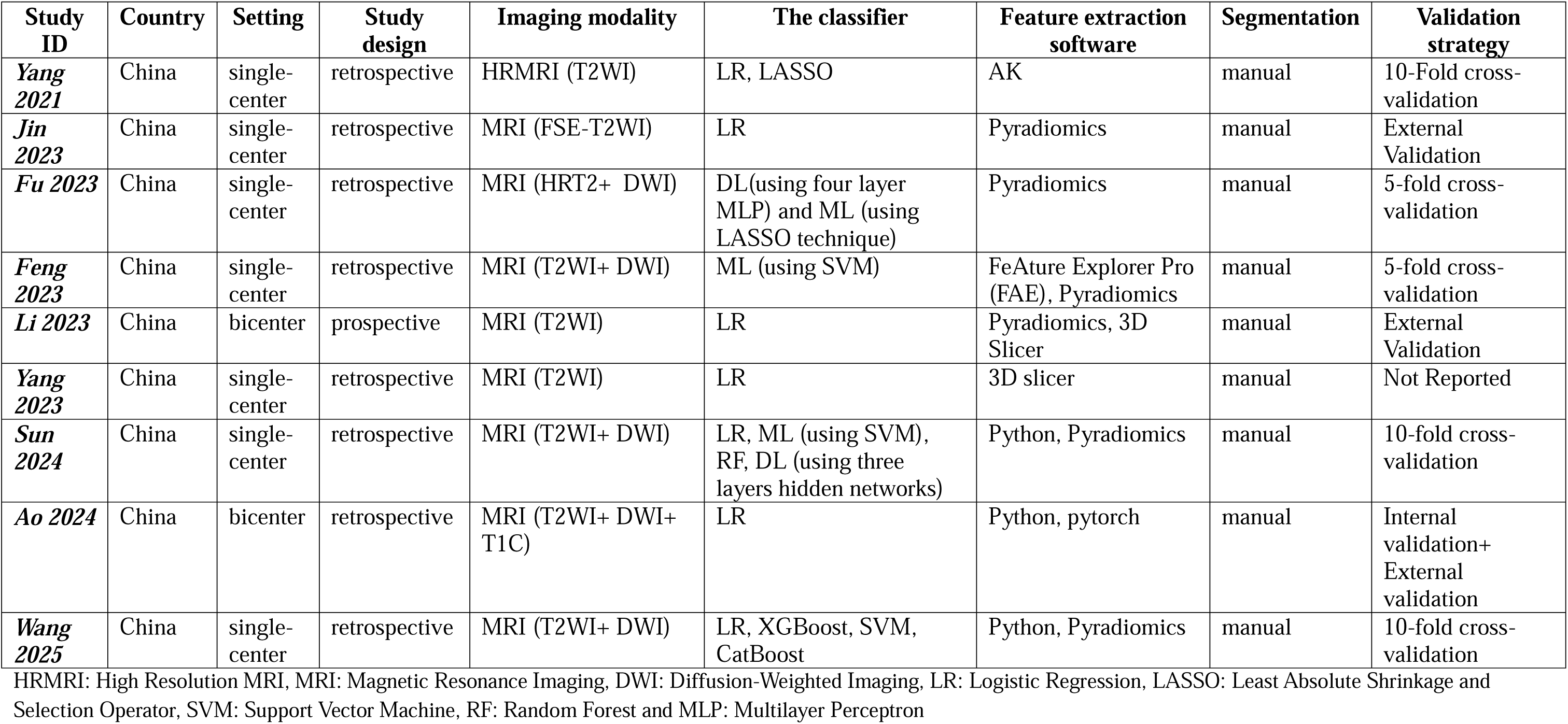
Summary of included studies.

| **Study ID** | **Country** | **Setting** | **Study design** | **Imaging modality** | **The classifier** | **Feature extraction software** | **Segmentation** | **Validation strategy** |
|---|---|---|---|---|---|---|---|---|
| ***Yang 2021*** | China | single-center | retrospective | HRMRI (T2WI) | LR, LASSO | AK | manual | 10-Fold cross-validation |
| ***Jin 2023*** | China | single-center | retrospective | MRI (FSE-T2WI) | LR | Pyradiomics | manual | External Validation |
| ***Fu 2023*** | China | single-center | retrospective | MRI (HRT2+ DWI) | DL(using four layer MLP) and ML (using LASSO technique) | Pyradiomics | manual | 5-fold cross-validation |
| ***Feng 2023*** | China | single-center | retrospective | MRI (T2WI+ DWI) | ML (using SVM) | FeAture Explorer Pro (FAE), Pyradiomics | manual | 5-fold cross-validation |
| ***Li 2023*** | China | bicenter | prospective | MRI (T2WI) | LR | Pyradiomics, 3D Slicer | manual | External Validation |
| ***Yang 2023*** | China | single-center | retrospective | MRI (T2WI) | LR | 3D slicer | manual | Not Reported |
| ***Sun 2024*** | China | single-center | retrospective | MRI (T2WI+ DWI) | LR, ML (using SVM), RF, DL (using three layers hidden networks) | Python, Pyradiomics | manual | 10-fold cross-validation |
| ***Ao 2024*** | China | bicenter | retrospective | MRI (T2WI+ DWI+ T1C) | LR | Python, pytorch | manual | Internal validation+ External validation |
| ***Wang 2025*** | China | single-center | retrospective | MRI (T2WI+ DWI) | LR, XGBoost, SVM, CatBoost | Python, Pyradiomics | manual | 10-fold cross-validation |

HRMRI: High Resolution MRI, MRI: Magnetic Resonance Imaging, DWI: Diffusion-Weighted Imaging, LR: Logistic Regression, LASSO: Least Absolute Shrinkage and Selection Operator, SVM: Support Vector Machine, RF: Random Forest and MLP: Multilayer Perceptron

### 3.2 Risk of bias assessment

Risk of bias assessment of the included studies was done using the QUADAS-2 tool. Assessment revealed that five studies (Jin et al. 2023 ^27^, Fu et al. 2023 ^28^, Feng 2023 ^29^, Sun et al. 2024 ^32^ and Ao et al. 2024 ^33^) showed low risk of bias. Moreover, the remaining four studies (Yang et al. 2021 ^26^, Li et al. 2023 ^27^, Yang et al. 2023 ^31^ and Wang et al. 2025 ^34^) demonstrated some concerns. Figure 2 shows the risk of bias graph of the included studies.

**Figure 2.**
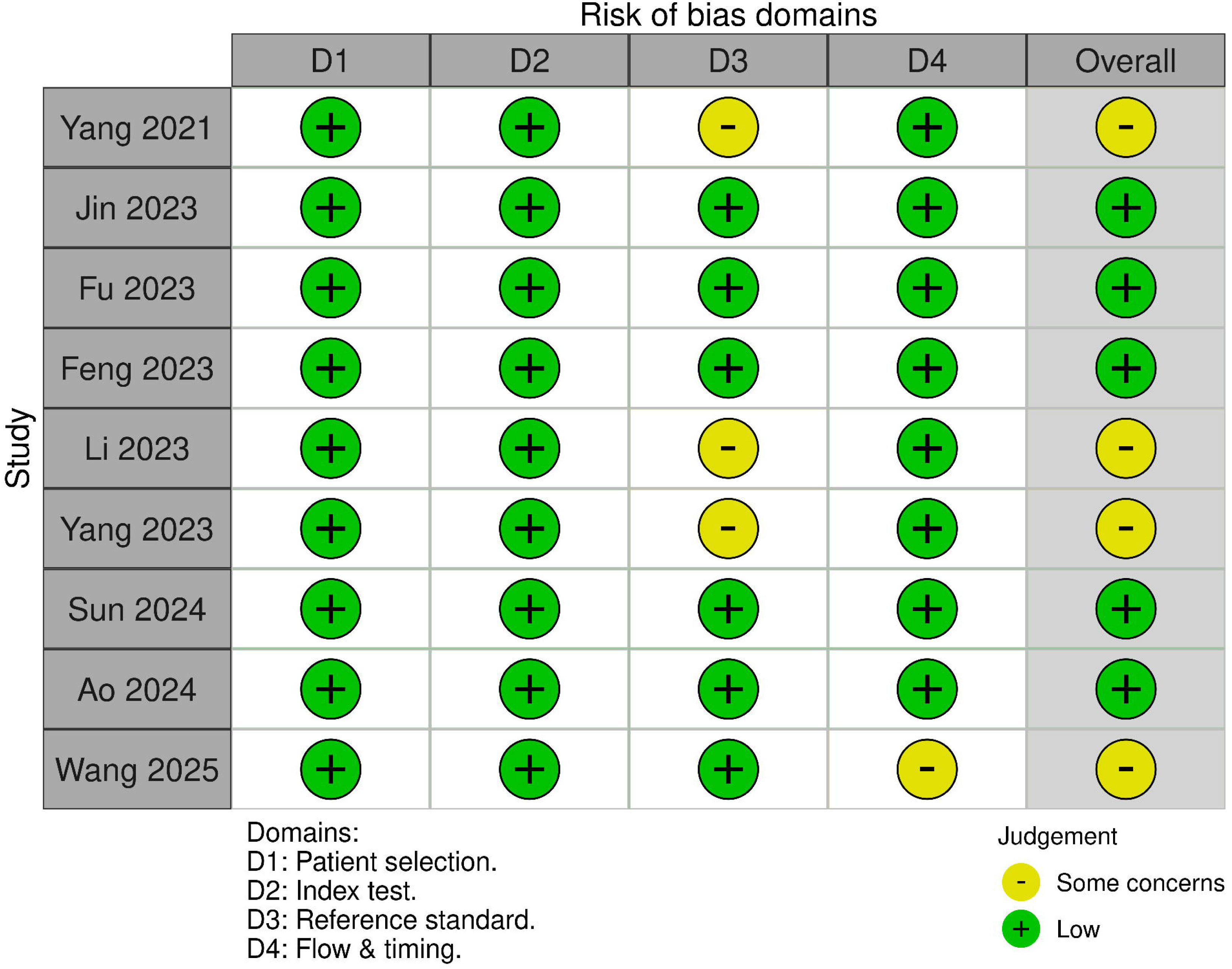
QUADAS-2 risk of bias and applicability assessment. Methodological quality of the included studies assessed using the QUADAS-2 tool across four domains: patient selection (D1), index test (D2), reference standard (D3), and flow and timing (D4). Green indicates low risk of bias and yellow indicates some concerns.

### 3.3 DTA pooled analysis

Across all AI cohorts, a meta-analysis using a bivariate model showed pooled sensitivity of 0.765 (95% CI = [0.714, 0.809]) and specificity of 0.79 (95% CI = [0.825, 0.75]), with an overall AUC of 0.845. Assessing between-studies heterogeneity using the Zhou and Dendukuri approach revealed very low heterogeneity (I^2^= 7.3%). (Figure 3)

**Figure 3.**
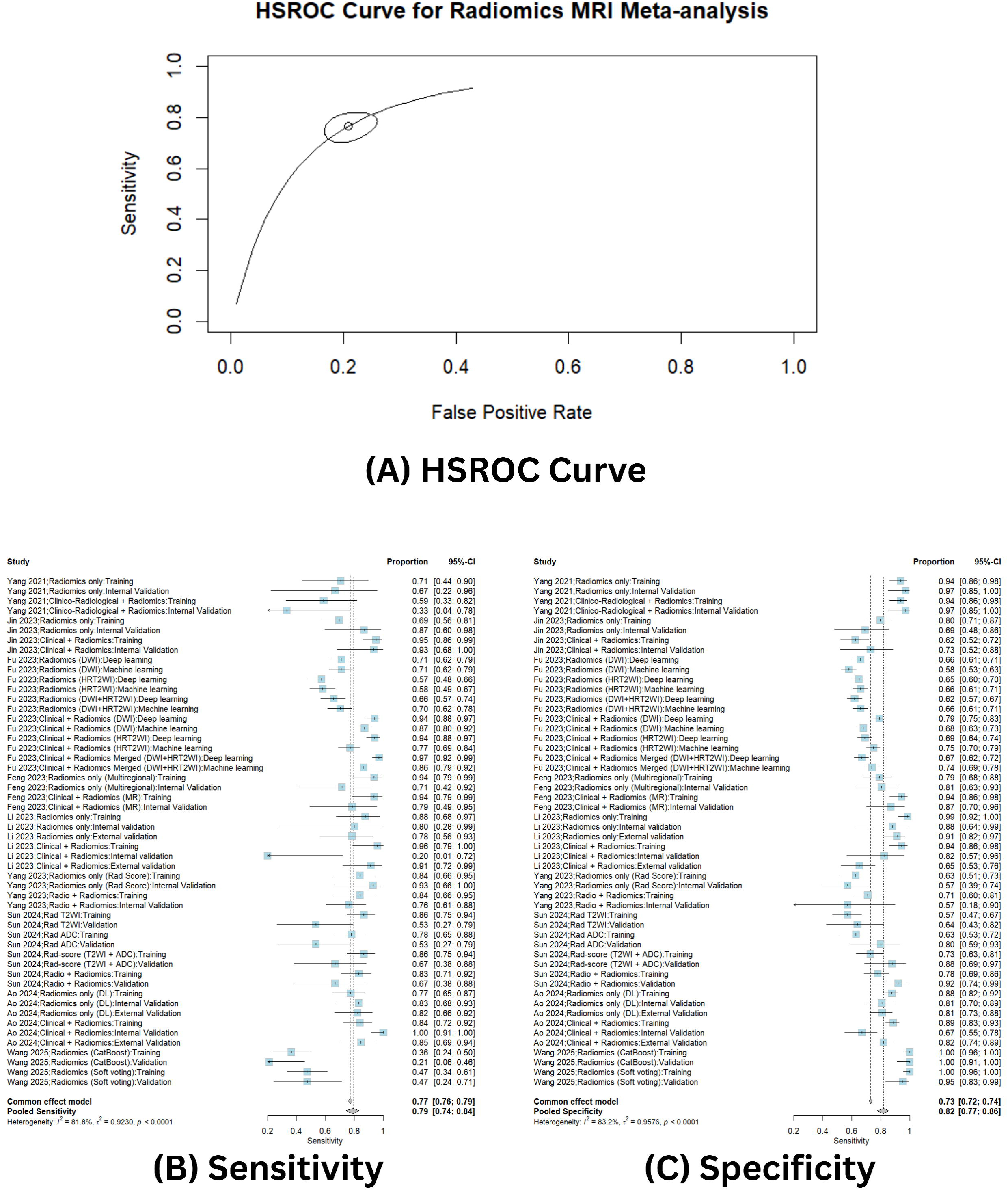
Diagnostic accuracy meta-analysis for development datasets (all AI cohorts): (A) HSROC curve generated from the bivariate random-effects model showing overall diagnostic performance of MRI-based radiomics models for detecting tumor deposits. (B) Forest plot of pooled sensitivity with 95% confidence intervals. (C) Forest plot of pooled specificity with 95% confidence intervals.

For validation datasets, pooled analysis showed that radiomics models showed a pooled AUC of 0.806, a sensitivity of 0.751 (95% CI = [0.681, 0.809]), and a specificity of 0.748 (95% CI = [0.708, 0.784]). Heterogeneity was very low (I^2^= 7.5%). (Figure 4)

**Figure 4.**
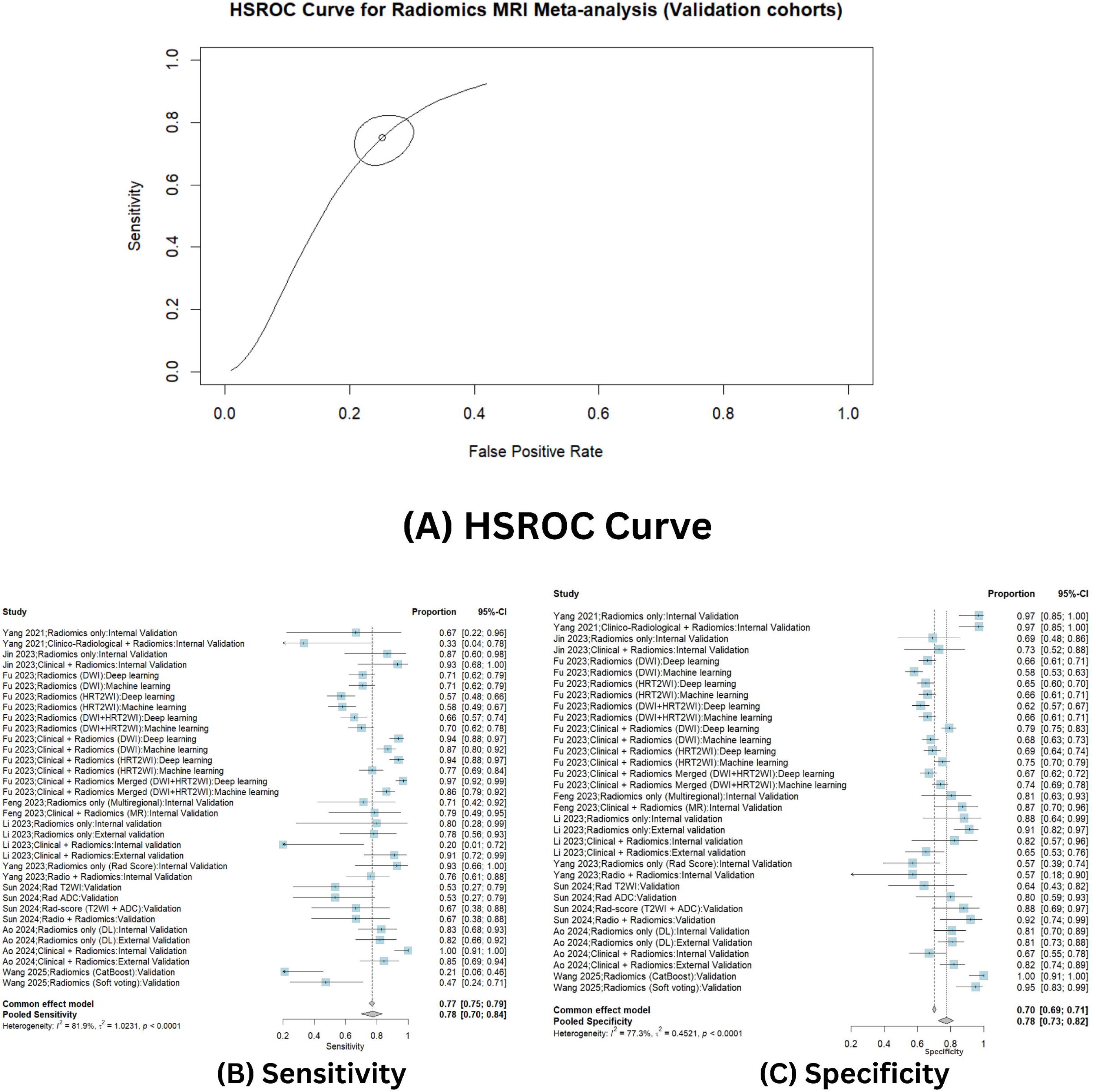
Diagnostic accuracy meta-analysis for validation datasets. (A) HSROC curve demonstrating pooled diagnostic performance in validation cohorts. (B) Forest plot of sensitivity estimates with 95% confidence intervals. (C) Forest plot of specificity estimates with 95% confidence intervals.

We performed predefined subgroup analyses to examine whether diagnostic performance varied by MRI input (T2WI vs T2WI+DWI) and by model design (radiomics alone vs radiomics combined with clinical/radiological variables). Detailed pooled estimates of sensitivity, specificity, AUC, and corresponding p-values for between-subgroup comparisons are summarized in Table S2 in the supplementary materials.

#### 3.3.1 MRI Input Subgroup Analysis (T2WI vs T2WI+DWI)

In subgroup analysis of pooled DTA, T2WI alone showed pooled sensitivity of 0.745 (95% CI: 0.625–0.837), compared with 0.697 (95% CI: 0.527–0.825) for combined T2WI+DWI. However, subgroup difference was not statistically significant (p = 0.607), indicating no clear evidence that adding DWI improves sensitivity compared with T2WI alone.

Pooled specificity showed an opposite pattern as T2WI+DWI achieved a higher specificity of 0.809 (95% CI: 0.707–0.881) compared with 0.737 (95% CI: 0.668–0.796) for T2WI alone. The difference was also not statistically significant (p = 0.192).

The pooled area under the curve (AUC) was slightly higher for T2WI+DWI (0.829) than for T2WI alone (0.796), suggesting a modest improvement in overall discriminatory performance with multi-sequence input. Between study heterogeneity in the bivariate model showed minimal heterogeneity (Zhou and Dendukuri approach; T2WI alone I² = 14.4% and T2WI+DWI I²= 28.3%). (Figures S1 and S2 in the supplementary materials show results of pooled DTA of this subgroup)

#### 3.3.2 Model Design Subgroup Analysis (Radiomics Only vs Radiomics + Clinical/Radiological Variables)

On comparison of pure radiomics models versus models that combined radiomics features plus clinical/radiological variables, there was a marked difference in sensitivity. The pooled sensitivity of radiomics-only models was 0.657 (95% CI: 0.593–0.715), whereas the pooled sensitivity of combined models was 0.842 (95% CI: 0.749–0.904). The absolute difference was −0.185, indicating substantially higher sensitivity in the combined models, and this difference was statistically significant (p < 0.001), suggesting that integrating clinical/radiological information meaningfully enhances TDs detection beyond radiomics alone.

Specificity was similar between groups. Radiomics-only models had a pooled specificity of 0.758 (95% CI: 0.689–0.816), while combined models had 0.753 (95% CI: 0.711–0.791). The absolute difference was +0.005, which was not statistically significant (p = 0.896).

The pooled AUC increased from 0.752 in radiomics-only models to 0.827 in radiomics plus clinical/radiological models, consistent with better overall discrimination when clinical/radiological context is integrated. Between study heterogeneity in the bivariate model showed minimal heterogeneity (Zhou and Dendukuri approach; Radiomics only I² = 11% and Radiomics combined I²= 7.3%). (Figures S3 and S4 in the supplementary materials show results of pooled diagnostic test accuracy of this subgroup)

#### 3.3.3. AI Model Subgroup Analysis (Deep Learning vs Machine Learning vs Logistic Regression)

We assessed the diagnostic performance according to the types of AI models used: deep learning (DL), conventional machine learning (ML), and logistic regression (LR). This was done separately for models based on only radiomics and for models that integrated radiomics plus clinical/radiological variables. All the numeric results, including pooled estimates with 95% CIs, pairwise comparison p-values, and other pertinent data for the analyses, are fully consolidated and presented in Table 2 and visualized in Figure 5.

**Figure 5.**
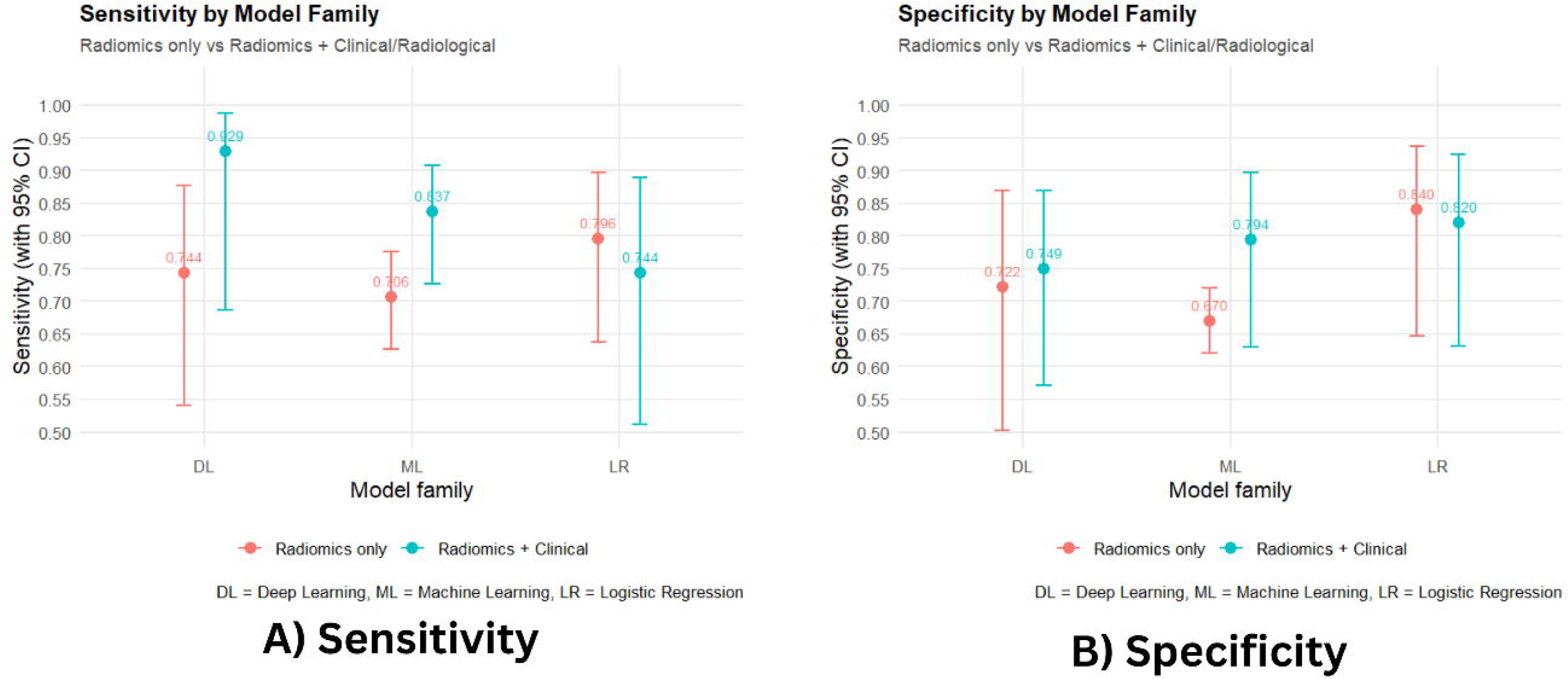
Subgroup DTA by AI model type: Diagnostic performance according to model family: deep learning (DL), machine learning (ML), and logistic regression (LR), stratified by model design (radiomics-only vs radiomics combined with clinical/radiological variables). (A) Sensitivity and (B) specificity with 95% confidence interval

**Table 2.** AI Model Subgroup Analysis (Deep Learning vs Machine Learning vs Logistic Regression)

| Comparison context | Pair (Arm1 vs Arm2) | Metric | Arm1 (value [95% CI]) | Arm2 (value [95% CI]) | p-value |
|---|---|---|---|---|---|
| *Within Radiomics only (DL vs ML vs LR)* | Rad only DL vs Rad only ML | Sensitivity | 0.744 [0.541–0.877] | 0.706 [0.627–0.775] | 0.685 |
|  |  | Specificity | 0.722 [0.502–0.870] | 0.670 [0.620–0.720] | 0.593 |
|  |  | AUC | 0.795 | 0.713 | — |
|  | Rad only DL vs Rad only LR | Sensitivity | 0.744 [0.541–0.877] | 0.796 [0.637–0.897] | 0.631 |
|  |  | Specificity | 0.722 [0.502–0.870] | 0.840 [0.647–0.937] | 0.324 |
|  |  | AUC | 0.795 | 0.872 | — |
|  | Rad only ML vs Rad only LR | Sensitivity | 0.706 [0.627–0.775] | 0.796 [0.637–0.897] | 0.238 |
|  |  | Specificity | 0.670 [0.620–0.720] | 0.840 [0.647–0.937] | **0.030**\* |
|  |  | AUC | 0.713 | 0.872 | — |
| *Within Radiomics + Clinical/Radiological (DL vs ML vs LR)* | Rad combined DL vs Rad combined ML | Sensitivity | 0.929 [0.687–0.987] | 0.837 [0.726–0.908] | 0.304 |
|  |  | Specificity | 0.749 [0.572–0.869] | 0.794 [0.630–0.897] | 0.659 |
|  |  | AUC | 0.883 | 0.882 | — |
|  | Rad combined DL vs Rad combined LR | Sensitivity | 0.929 [0.687–0.987] | 0.744 [0.512–0.889] | 0.132 |
|  |  | Specificity | 0.749 [0.572–0.869] | 0.820 [0.631–0.924] | 0.505 |
|  |  | AUC | 0.883 | 0.851 | — |
|  | Rad combined ML vs Rad combined LR | Sensitivity | 0.837 [0.726–0.908] | 0.744 [0.512–0.889] | 0.384 |
|  |  | Specificity | 0.794 [0.630–0.897] | 0.820 [0.631–0.924] | 0.797 |
|  |  | AUC | 0.882 | 0.851 | — |
| *Effect of adding Clinical/Radiological features (within same model type)* | Rad only DL vs Rad combined DL | Sensitivity | 0.744 [0.541–0.877] | 0.929 [0.687–0.987] | 0.107 |
|  |  | Specificity | 0.722 [0.502–0.870] | 0.749 [0.572–0.869] | 0.823 |
|  |  | AUC | 0.795 | 0.883 | — |
|  | Rad only ML vs Rad combined ML | Sensitivity | 0.706 [0.627–0.775] | 0.837 [0.726–0.908] | **0.029**\* |
|  |  | Specificity | 0.670 [0.620–0.720] | 0.794 [0.630–0.897] | 0.088 |
|  |  | AUC | 0.713 | 0.882 | — |
|  | Rad only LR vs Rad combined LR | Sensitivity | 0.796 [0.637–0.897] | 0.744 [0.512–0.889] | 0.656 |
|  |  | Specificity | 0.840 [0.647–0.937] | 0.820 [0.631–0.924] | 0.849 |
|  |  | AUC | 0.872 | 0.851 | — |

\*: Statistically significant, DL: Deep learning, ML: Machine Learning, LR: Logistic Regression

For radiomics-only models, pooled sensitivity was 0.744 (95% CI: 0.541–0.877) for DL, 0.706 (95% CI: 0.627–0.775) for ML, and 0.796 (95% CI: 0.637–0.897) for LR. These differences in sensitivity between DL, ML, and LR were not statistically significant (all p>0.05). Pooled specificity was 0.722 (95% CI: 0.502–0.870) for DL, 0.670 (95% CI: 0.620–0.720) for ML, and 0.840 (95% CI: 0.647– 0.937) for LR. Logistic regression showed significantly higher pooled specificity than ML (p=0.030), whereas the differences between DL and ML and between DL and LR were not statistically significant (p>0.05). The pooled AUCs for radiomics-only models were 0.795 (DL), 0.713 (ML), and 0.872 (LR), indicating that LR tended to have stronger overall discriminative ability than ML in radiomics only setting. (Figures S5-S7 in the supplementary materials)

For radiomics models combined with clinical/radiological variables, pooled sensitivity was 0.929 (95% CI: 0.687–0.987) for DL, 0.837 (95% CI: 0.726–0.908) for ML, and 0.744 (95% CI: 0.512– 0.889) for LR. Pooled specificity was 0.749 (95% CI: 0.572–0.869) for DL, 0.794 (95% CI: 0.630– 0.897) for ML, and 0.820 (95% CI: 0.631–0.924) for LR. None of the pairwise differences in sensitivity or specificity between DL, ML, and LR within this combined category were statistically significant (all p>0.05). The pooled AUCs were uniformly high ـــ 0.883 (DL), 0.882 (ML), and 0.851 (LR) ـــ suggesting that once clinical/radiological context is added, all three model types achieve broadly comparable overall discrimination. (Figures S8-S10 in the supplementary materials)

## 4 DISCUSSIONS

Treatment strategy for RC has shown a paradigm shift after introducing the concept of total mesorectal excision (TME) by Heald ^35^ which later was combined with the neoadjuvant chemoradiotherapy ^36^. These advances continued with the incorporation of pelvic MRI with a tailored surgical resection plane based on preoperative MRI enabled reduction of circumferential resection margin involvement.

Fryback and Thornbury ^37^ employed a hierarchical model to evaluate the added value of newly introduced technology in clinical practice, which testified MRI as a validated imaging modality for RC evaluation ^38^. A further quality improvement in RC evaluation was achieved with implementation of the structured reporting or proformas, with an improvement of 78% in report completeness ^39^. These proformas incorporated entities of TDs, extramural vascular invasion (EMVI), and perineural invasion (PNI), which showed a significantly poorer prognosis ^8,40^ even greater than tumor and nodal stage ^41^.

TDs are mesorectal nodules of malignant cells of uncertain origin discontinuous from the primary tumor ^40^. Pelvic MRI shows TDs as irregular margins nodules along the vessels, even interrupting their course, however, should be differentiated from lymph nodes ^41^.

Radiomics and radiogenomics have been fields of innovative research in the last years in RC. Radiomics implies AI model to mine a vast number of qualitative and quantitative features that cannot be captured from routine radiographs by human eye ^42^. Several research have been studied the accuracy of MRI-based radiomics in prediction of pathological complete response, restaging MRI, genetic profiles of tumors (radiogenomics), and prediction of distant metastases ^43^.

Despite promising data reported with high performance metrics such as sensitivity, specificity, accuracy, and AUC, however, several challenges stand against implementation such as small sample dataset, algorithm generalizability, and integration in clinical practice ^44^.

To the best of our knowledge this study is the first DTA meta-analysis studying the role of MRI-based radiomics in prediction of TDs in RC patients. In this study, MRI-based radiomics models demonstrate reliable detection of TDs in RC, with reproducible discrimination across included studies which are largely persisting in validation subsets. These findings are clinically relevant, given the high ambiguity of tumor deposits with small nodes, vessels, fibrosis, and post-inflammatory changes. The delineation of subgroups affords clinical insight.

First, expanding the image data input from high-resolution T2WI to include DWI did not result in consistent improvement in TDs detection, reinforcing the notion that TDs detection is not primarily limited by the visibility of TDs on one of the two sequences, but rather by the ambiguity of the TDs and other similar structures and the inherent uncertainty. This aligns with contemporary rectal MRI practice, where high-quality T2WI remains an essential component for initial RC detection and staging, while DWI is used only when the image quality and interpretative context support its inclusion ^45^.

Second and most consequential for clinical translation the models that combine radiomics with clinical/radiological parameters seem to have achieved significantly better case detection with no commensurate increase in false positive detection. This is biologically understandable, as the TDs detection probability is heavily dependent on tumor phenotype and staging context (e.g., extramural spread, vascular invasion proxies, mesorectal risk) features that are not captured by texture alone.

Overall, a deployment-ready TDs model should be context-fused by design (radiomics plus structured radiological/clinical predictors), built on high-resolution oblique axial T2W MRI as mandatory core input, with DWI/ADC included as an optional auxiliary stream primarily to stabilize specificity, using IBSI-standardized feature extraction and transparent reporting ^46,47^.

The fact that no consistent "winner" tends to emerge across algorithm families once clinical/radiological context is factored in supports a core assumption for TDs detection, model formulation and validation approach take precedence over classifier choice. This reinforces the findings from clinical prediction studies, where apparent improvements from machine learning over logistic regression often diminish in value in pragmatic validation and full disclosure ^48^.

The recurrent signal of high sensitivity with deep learning should also be considered. A possible explanation is that DL feature learning can capture structured/contextual information, such as irregularities in nodules, patterns at the mesorectal invasion, and relationships to the primary tumor, which radiomic features may lose through their handcrafted process, thereby aiding in case-finding ^49^.

The opposite argument is that sensitivity is the metric most susceptible to class imbalance, threshold selection, and domain shift. DL models, especially in data-limited, single- or few-center settings typical of TD datasets, are prone to shortcut learning and calibration drift, leading to ‘optimistic’ sensitivity that does not generalize ^50^. Thus, if the intended clinical role is “flagging” (maximizing detection), DL is a justifiable contender only in the context of a multimodal fusion framework and only post locked-threshold external validation plus calibration and workflow assessments; if not, then regularized regression or gradient-boosted models based on fused radiomics plus structured predictors may provide similar discrimination with better stability and interpretability, and will be more readily implementable in the near-term.

Although the results remain encouraging, certain limitations and considerations need to be noted. The first being that the number of studies is still small, and all the studies being examined are from China. This limits the generalizability of the findings to other countries and health care systems. Secondly, as most studies are single-center, retrospective studies, they are more prone to selection bias and may exhibit greater model performance inappropriately, compared to prospective multicenter studies. The third consideration is the lack of uniformity across studies in MRI acquisition, segmentation, radiomic feature extraction software, and modeling methodologies. This could compromise the stability and reproducibility of the features. The fourth limitation is that several studies relied on internal validation, which most likely limited their external validation. This could potentially lead to great overestimates of the study’s diagnostic inconsistency. Lastly, the reports on the studies of different AI systems and sandwiches were inconsistent and not standardized. Therefore, the need for more studies is great in the area of utilizing high-grade, validated radiomics, multicenter, prospective studies in greater numbers.

Future research should prioritize large multicenter prospective studies to improve the generalizability and clinical reliability of AI-based MRI radiomics models for tumor deposit detection. Standardization of imaging acquisition parameters, segmentation methods, and radiomics feature extraction—such as adherence to the Image Biomarker Standardisation Initiative (IBSI) framework—will be important for improving reproducibility across institutions ^51^. Moreover, future models should integrate radiomic features with clinical, pathological, and conventional imaging variables, as multimodal approaches may better capture tumor biology and improve predictive performance. Finally, rigorous external validation and clinical impact studies are needed to determine whether these models can meaningfully improve preoperative staging and support treatment decision-making in routine clinical practice.

## 5 CONCLUSION

MRI-based radiomics shows good performance for the preoperative identification of tumor deposits in rectal cancer, exhibiting consistent sensitivity and specificity across all studies and validation datasets. Case detection is enhanced when radiomics is integrated with other clinical or radiological elements compared to radiomics in isolation, demonstrating the value of multimodal prediction approaches. Although high-resolution T2-weighted MRI primarily maintains the fundamental imaging input, the addition of diffusion-weighted imaging mildly increases specificity and overall discriminating performance. In conclusion, although AI-based MRI radiomics provides a promising non-invasive method for preoperative risk stratification and rectal cancer treatment planning, wider external validation and standardized radiomics procedures are critical for clinical use.

## Data Availability

All data produced in the present study are available upon reasonable request to the authors

## List of abbreviations

AI: Artificial Intelligence
AUC: Area Under the Curve
CI: Confidence Interval
CRC: Colorectal Cancer
CT: Computed Tomography
DL: Deep Learning
DTA: Diagnostic Test Accuracy
DWI: Diffusion Weighted Imaging
EMVI: Extramural Vascular Invasion
ENTDs: Extranodal Tumor Deposits
FAE: FeAture Explorer Pro
FN: False Negative
FP: False Positive
HSROC: Hierarchical Summary Receiver Operating Characteristic
IBSI: Image Biomarker Standardisation Initiative
LR: Logistic Regression
ML: Machine Learning
MLP: Multilayer Perceptron
MRI: Magnetic Resonance Imaging
PNI: Perineural Invasion
PRISMA-DTA: Preferred Reporting Items for Systematic Reviews and Meta-Analyses of Diagnostic Test Accuracy Studies
QUADAS-2: Quality Assessment of Diagnostic Accuracy Studies-2
RC: Rectal Cancer
RF: Random Forest
ROC: Receiver Operating Characteristic
SVM: Support Vector Machine
T1C: Contrast-Enhanced T1-Weighted Imaging
T2WI: T2-Weighted Imaging
TDs: Tumor Deposits
TME: Total Mesorectal Excision
TN: True Negative
TNM: Tumor Node Metastasis
TP: True Positive

