## Supplementary materials for "AI-Based MRI Radiomics for Preoperative Detection of Tumor Deposits in Rectal Cancer: A Bivariate Diagnostic Accuracy Meta-analysis"

**Supplementary Tables**

**Figure S1**. PRISMA-DTA Checklist

| **Section/topic** | **#** | **PRISMA-DTA Checklist Item** | **Reported on page #** |
| --- | --- | --- | --- |
| **TITLE / ABSTRACT** | | |  |
| Title | 1 | Identify the report as a systematic review (+/- meta-analysis) of diagnostic test accuracy (DTA) studies. | 1 |
| Abstract | 2 | Abstract: See PRISMA-DTA for abstracts. | 3-4 |
| **INTRODUCTION** | | |  |
| Rationale | 3 | Describe the rationale for the review in the context of what is already known. | 5-6 |
| Clinical role of index test | D1 | State the scientific and clinical background, including the intended use and clinical role of the index test, and if applicable, the rationale for minimally acceptable test accuracy (or minimum difference in accuracy for comparative design). | 5-6 |
| Objectives | 4 | Provide an explicit statement of question(s) being addressed in terms of participants, index test(s), and target condition(s). | 6 |
| **METHODS** | | |  |
| Protocol and registration | 5 | Indicate if a review protocol exists, if and where it can be accessed (e.g., Web address), and, if available, provide registration information including registration number. | 6 |
| Eligibility criteria | 6 | Specify study characteristics (participants, setting, index test(s), reference standard(s), target condition(s), and study design) and report characteristics (e.g., years considered, language, publication status) used as criteria for eligibility, giving rationale. | 7 |
| Information sources | 7 | Describe all information sources (e.g., databases with dates of coverage, contact with study authors to identify additional studies) in the search and date last searched. | 6 |
| Search | 8 | Present full search strategies for all electronic databases and other sources searched, including any limits used, such that they could be repeated. | 6 |
| Study selection | 9 | State the process for selecting studies (i.e., screening, eligibility, included in systematic review, and, if applicable, included in the meta-analysis). | 7-8 |
| Data collection process | 10 | Describe method of data extraction from reports (e.g., piloted forms, independently, in duplicate) and any processes for obtaining and confirming data from investigators. | 8 |
| Definitions for data extraction | 11 | Provide definitions used in data extraction and classifications of target condition(s), index test(s), reference standard(s) and other characteristics (e.g. study design, clinical setting). | 8 |
| Risk of bias and applicability | 12 | Describe methods used for assessing risk of bias in individual studies and concerns regarding the applicability to the review question. | 8 |
| Diagnostic accuracy measures | 13 | State the principal diagnostic accuracy measure(s) reported (e.g. sensitivity, specificity) and state the unit of assessment (e.g. per-patient, per-lesion). | 8-9 |
| Synthesis of results | 14 | Describe methods of handling data, combining results of studies and describing variability between studies. This could include, but is not limited to: a) handling of multiple definitions of target condition. b) handling of multiple thresholds of test positivity, c) handling multiple index test readers, d) handling of indeterminate test results, e) grouping and comparing tests, f) handling of different reference standards | 8-9 |

**Table S2.** Subgroup analyses according to MRI modality and according to model design

| **Comparison** | **Metric** | **Arm 1 (value [95% CI])** | **Arm 2 (value [95% CI])** | **p-value** |
| --- | --- | --- | --- | --- |
| ***T2WI vs T2WI+DWI*** | Sensitivity | T2WI: 0.745 [0.625–0.837] | T2WI+DWI: 0.697 [0.527–0.825] | 0.607 |
|  | Specificity | T2WI: 0.737 [0.668–0.796] | T2WI+DWI: 0.809 [0.707–0.881] | 0.192 |
|  | AUC | T2WI: 0.796 | T2WI+DWI: 0.829 | — |
| ***Radiomics only vs Radiomics + Clinical/Rad*** | Sensitivity | Radiomics only: 0.657 [0.593–0.715] | Radiomics + Clinical/Rad: 0.842 [0.749–0.904] | **<0.001*** |
|  | Specificity | Radiomics only: 0.758 [0.689–0.816] | Radiomics + Clinical/Rad: 0.753 [0.711–0.791] | 0.896 |
|  | AUC | Radiomics only: 0.752 | Radiomics + Clinical/Rad: 0.827 | — |

***** : statistically significant

**Supplementary Figures**

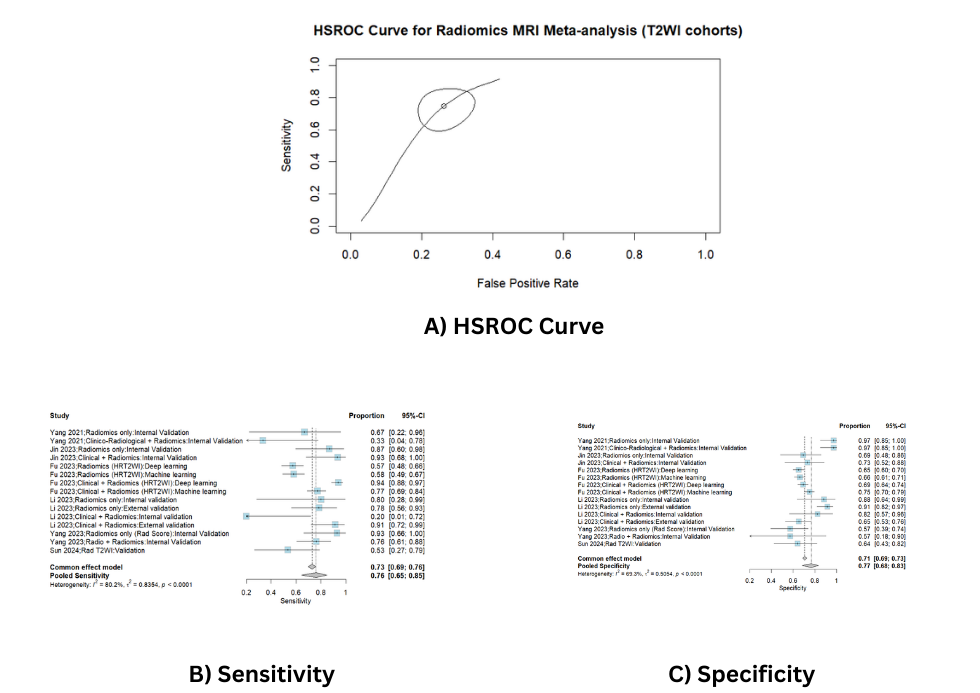

**Figure S1.** Pooled analysis of T2WI Cohorts; A) HSROC curve, B) Sensitivity forest plot and C) Specificity forest plot

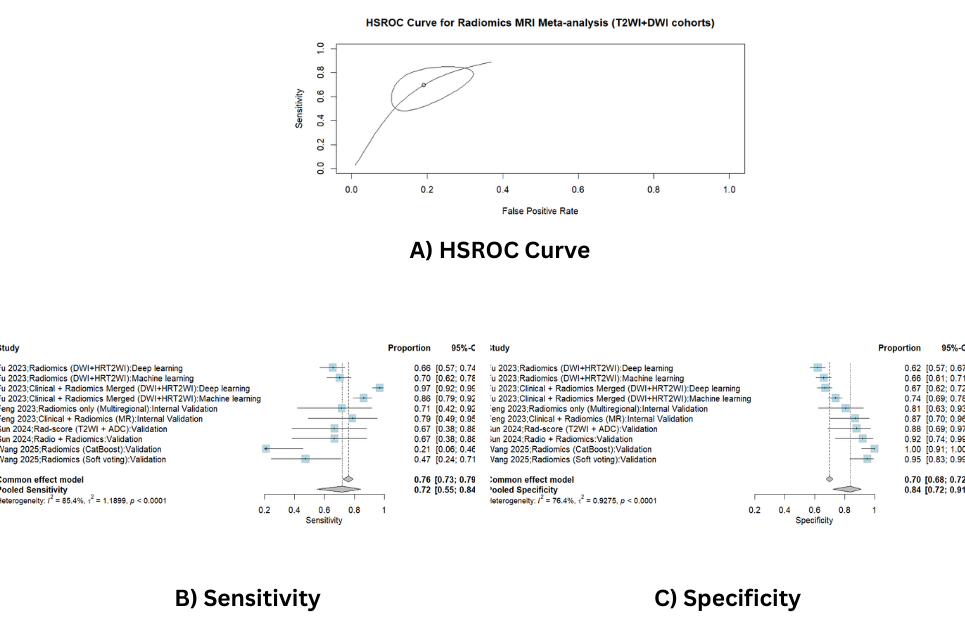

**Figure S2.** Pooled analysis of T2WI+DWI Cohorts; A) HSROC curve, B) Sensitivity forest plot and C) Specificity forest plot

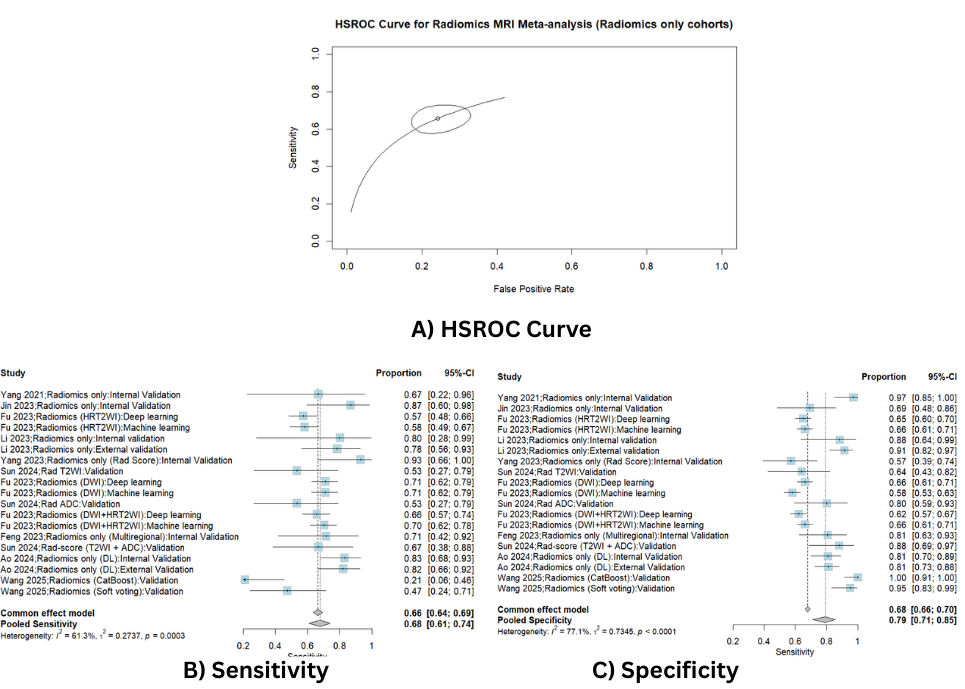

**Figure S3.** Pooled analysis of Radiomics only Cohorts; A) HSROC curve, B) Sensitivity forest plot and C) Specificity forest plot

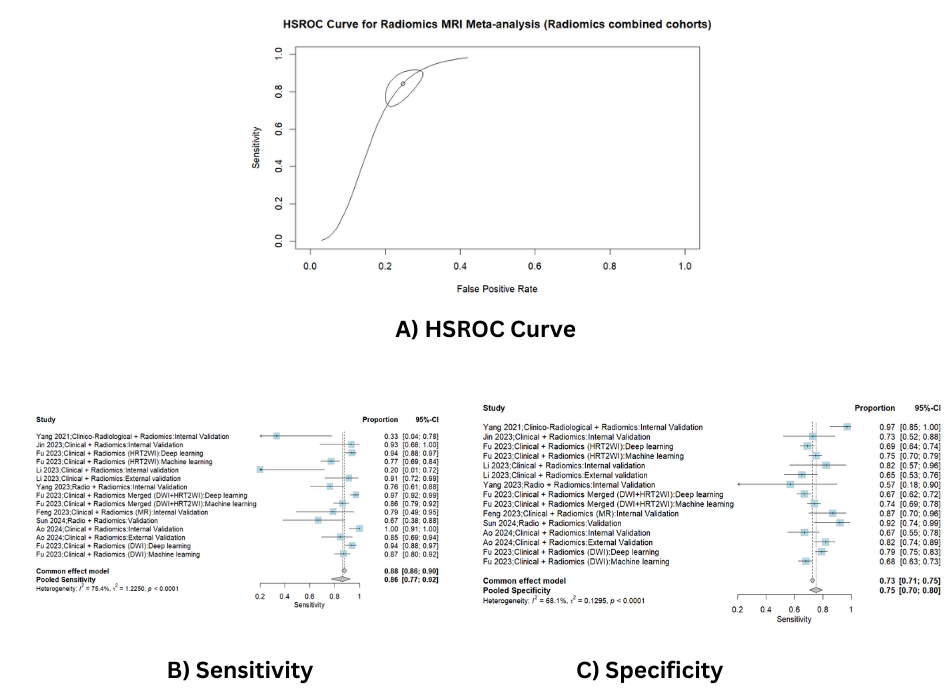

**Figure S4.** Pooled analysis of Radiomics combined with clinical or radiological parameters Cohorts; A) HSROC curve, B) Sensitivity forest plot and C) Specificity forest plot

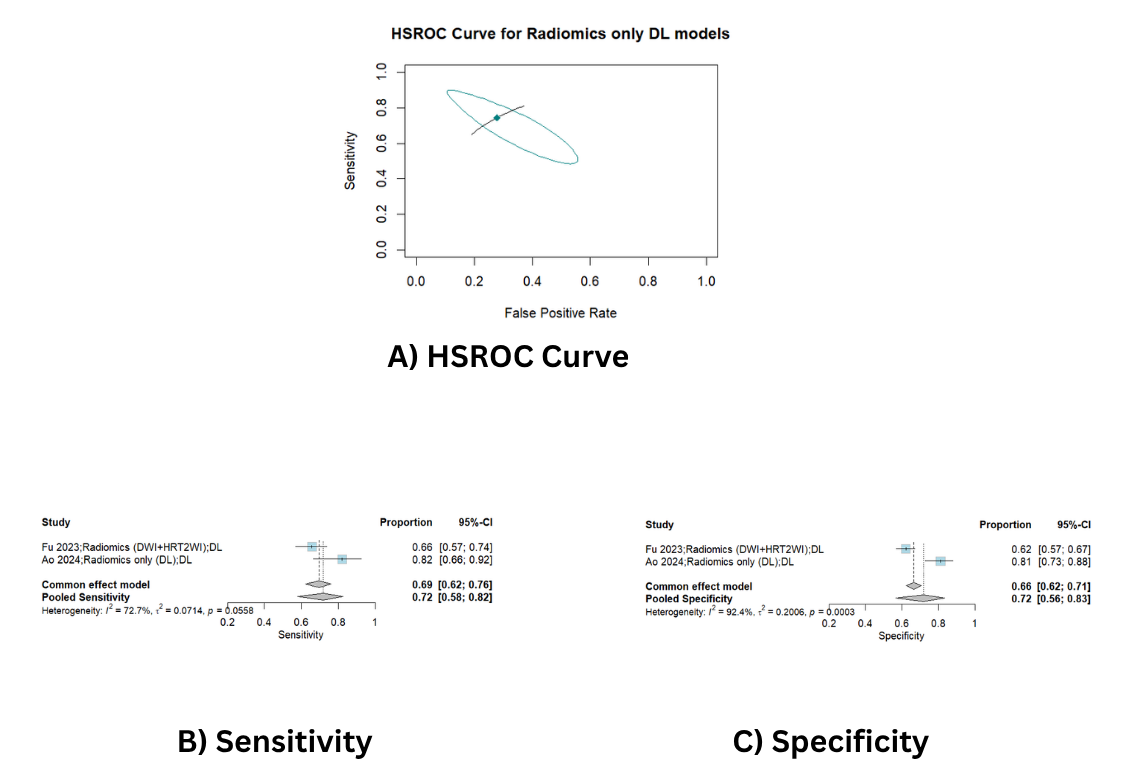

**Figure S5.** Pooled analysis of Radiomics only Deep Learning (DL) Cohorts; A) HSROC curve, B) Sensitivity forest plot and C) Specificity forest plot

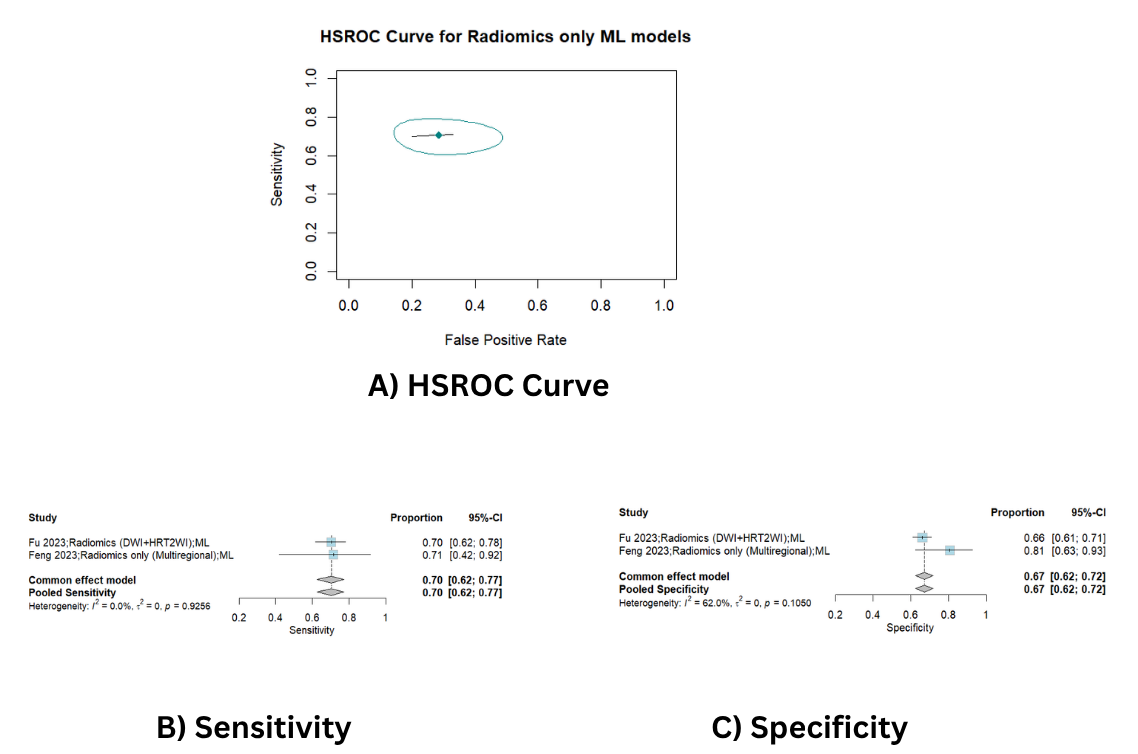

**Figure S6.** Pooled analysis of Radiomics only Machine Learning (ML) Cohorts; A) HSROC curve, B) Sensitivity forest plot and C) Specificity forest plot

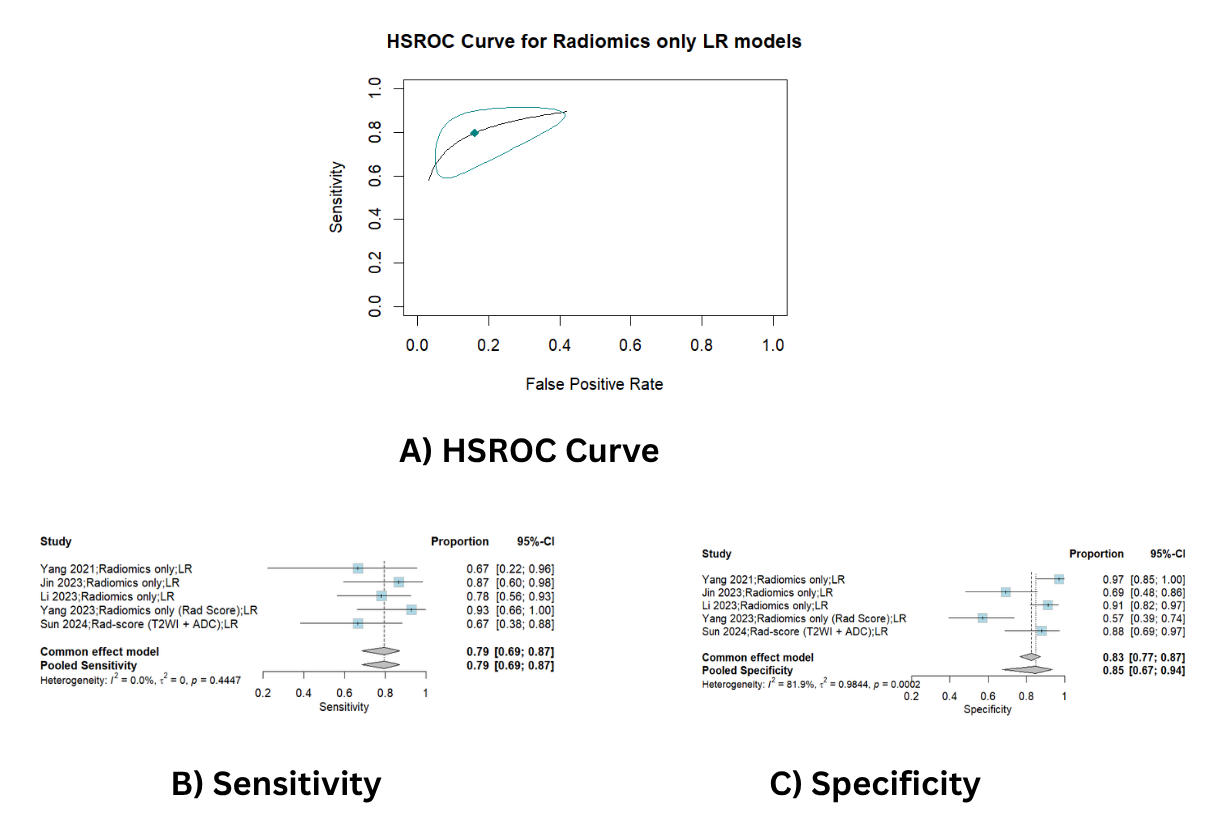

**Figure S7.** Pooled analysis of Radiomics only Logistic Regression (LR) Cohorts; A) HSROC curve, B) Sensitivity forest plot and C) Specificity forest plot

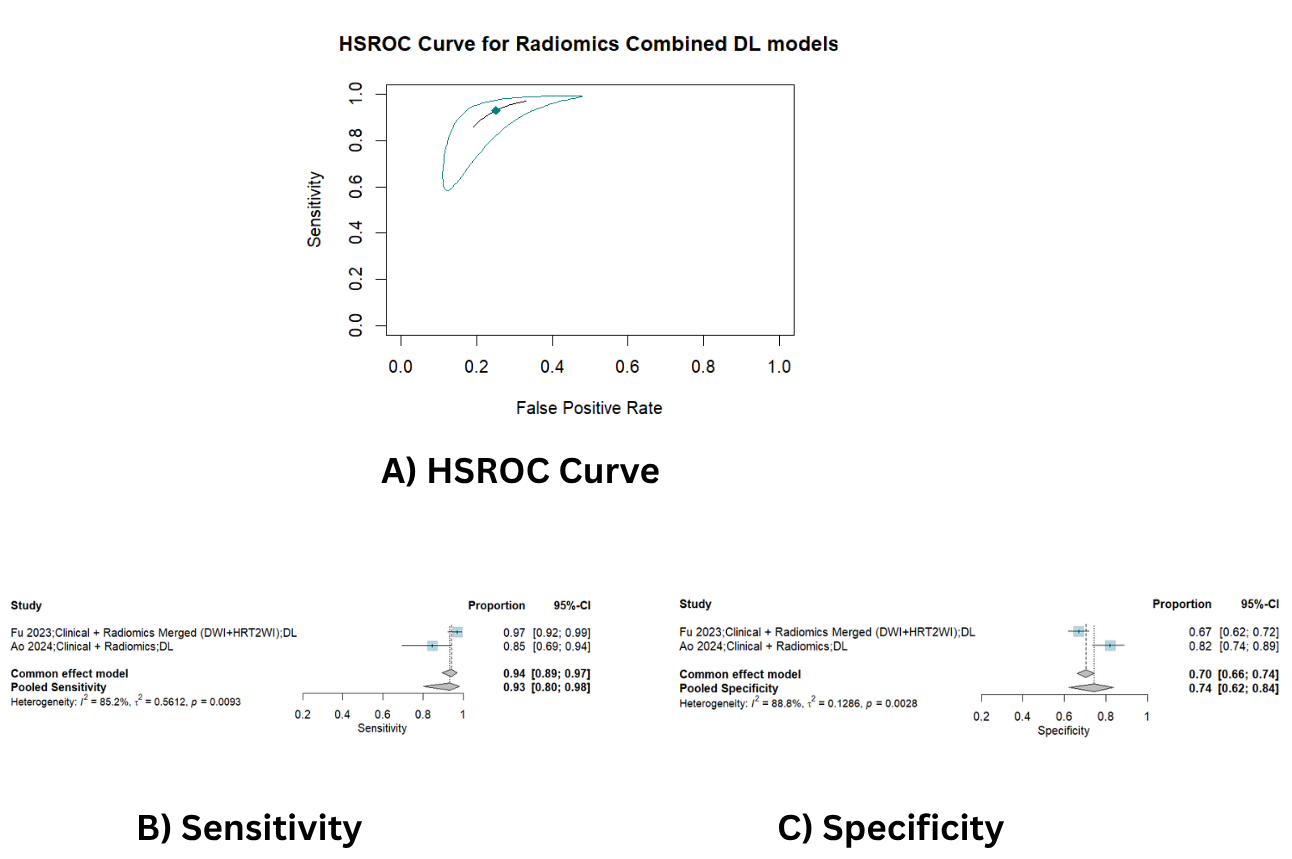

**Figure S8.** Pooled analysis of Radiomics combined with clinical or radiological parameters Deep Learning (DL) Cohorts; A) HSROC curve, B) Sensitivity forest plot and C) Specificity forest plot

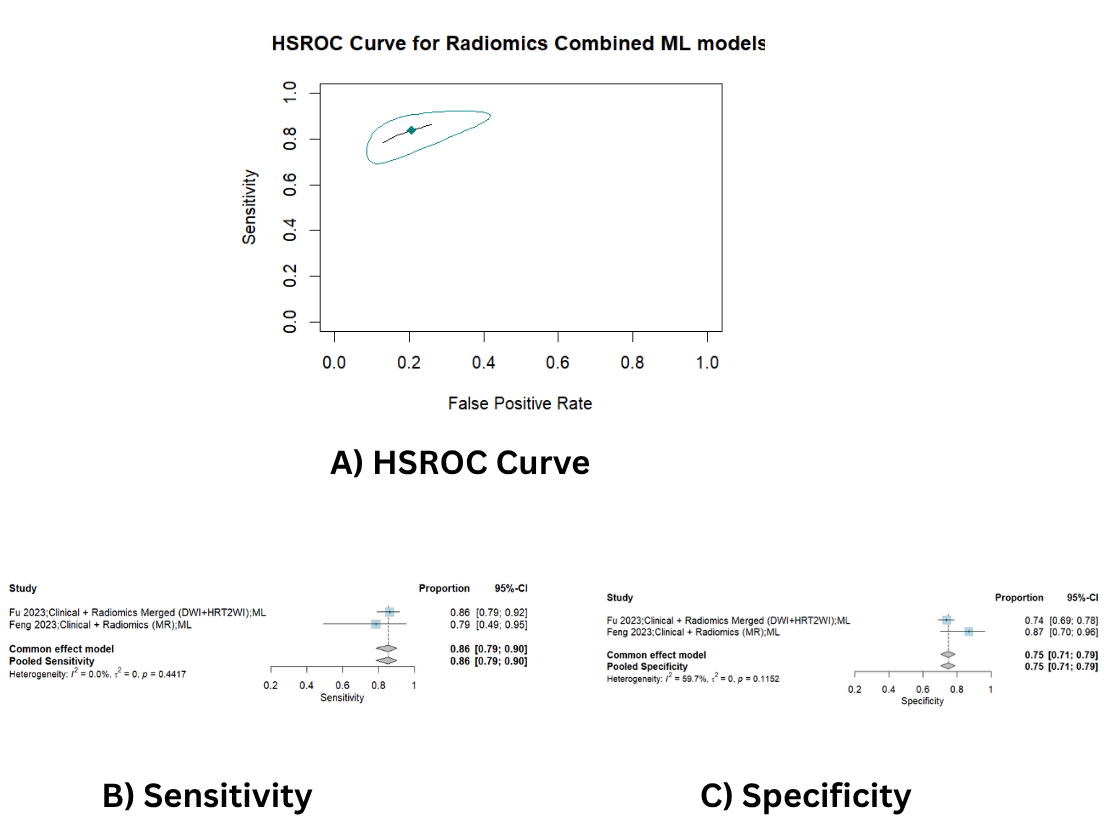

**Figure S9.** Pooled analysis of Radiomics combined with clinical or radiological parameters Machine Learning (ML) Cohorts; A) HSROC curve, B) Sensitivity forest plot and C) Specificity forest plot

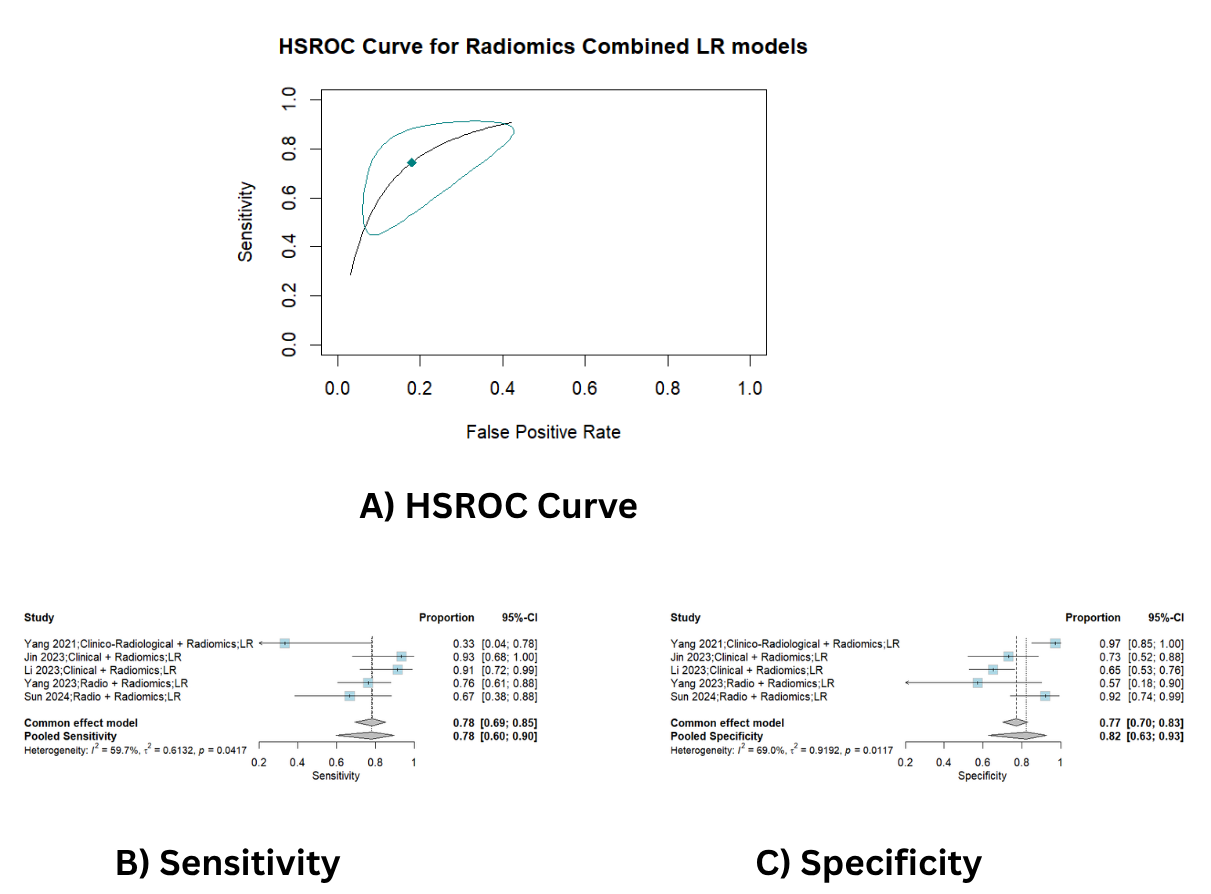

**Figure S10.** Pooled analysis of Radiomics combined with clinical or radiological parameters Logistic Regression (LR) Cohorts; A) HSROC curve, B) Sensitivity forest plot and C) Specificity forest plot
